# COVID-19 Propagation and Mortality in a Two-Part Population

**DOI:** 10.1101/2020.05.17.20104356

**Authors:** Malcolm. W. McGeoch, Julie E. M. McGeoch

**Affiliations:** PLEX Corporation, 275 Martine St., Suite 100, Fall River, MA 02723, USA; Dept. of Molecular and Cellular Biology, Harvard University, 52 Oxford St., Cambridge MA 02138, USA

## Abstract

There has recently emerged a striking consistency to the mortality from SARS-CoV-2, [1,2,3] as a fraction of population, across many nations. We have constructed a model for the spread of the virus that reproduces this phenomenon via inclusion of two (or more) categories of susceptibility to the virus. In the simplest case, the population is given a smaller fraction of 10-20% with higher susceptibility and the balance of 80-90% with lower susceptibility. Susceptibility is taken to include the level of immunity to the virus combined with the societal circumstances of certain smaller groups within a population. This is programmed numerically by considering a realistic random rate of contacts, together with an assumed constant viral genome. The remaining major variable is the societal response of nations to the outbreak, with earlier or later application of various degrees of lockdown, tracing and sanitation. China, South Korea and other nations, including Germany, have stopped or greatly slowed the spread of the disease before it could run its course through a whole population. Using this model the extent of progress toward herd immunity is discussed, with an in-principle estimate of the remaining toll to be experienced.

## 1 Introduction

The last thing the first author expected was to be applying his experience in modeling atomic and molecular processes in gas lasers to a sudden affliction that has within 100 days affected all of humanity. In these lasers there is usually a very large number of different states that can interact in different ways – complexity is the rule. The statistics of light emitted by lasers depends upon an initial growth from a single photon into a very powerful beam, an example of the process of exponential growth, which has become too familiar to us in the course of this disease. The puzzle that gave rise to the present calculation gradually developed over the most recent 20 days (up to 12^th^ May, 2020) during which the fractional death for many nations [4], plotted systematically by the second author, in Figure 1, converged on 0.05%. This could not be explained easily, because the mortality in diagnosed cases was in some nations very high: from 5 to 15%. If the disease was working its way through a uniform population of people identically susceptible to the disease, and propagation of the disease had not been suppressed via a change in behavior, then different countries would be exhibiting vastly different fractional mortality. This is because the time for cases to multiply by 2.7 times (one E-fold, discussed below) was measured from the traces in Figure 1 to be only 3.1 ± 0.5 days, which is much shorter than the spacing of different nations’ growth curves by 20 days at the same fractional point. There ought to have been orders of magnitude different fractional deaths at a given time, and many nations ought by now to have exceeded the observed 0.05% in Figure 1. On the other hand, had nations taken action to reduce gathering size and improve hygiene, then the differences between national approaches would again have stood out much more starkly than the constant result of Figure 1. The fact that several nations stand out by having much lower mortality, indicating effective mitigation, does not explain the constancy of mortality in the higher group.

**Figure 1.**
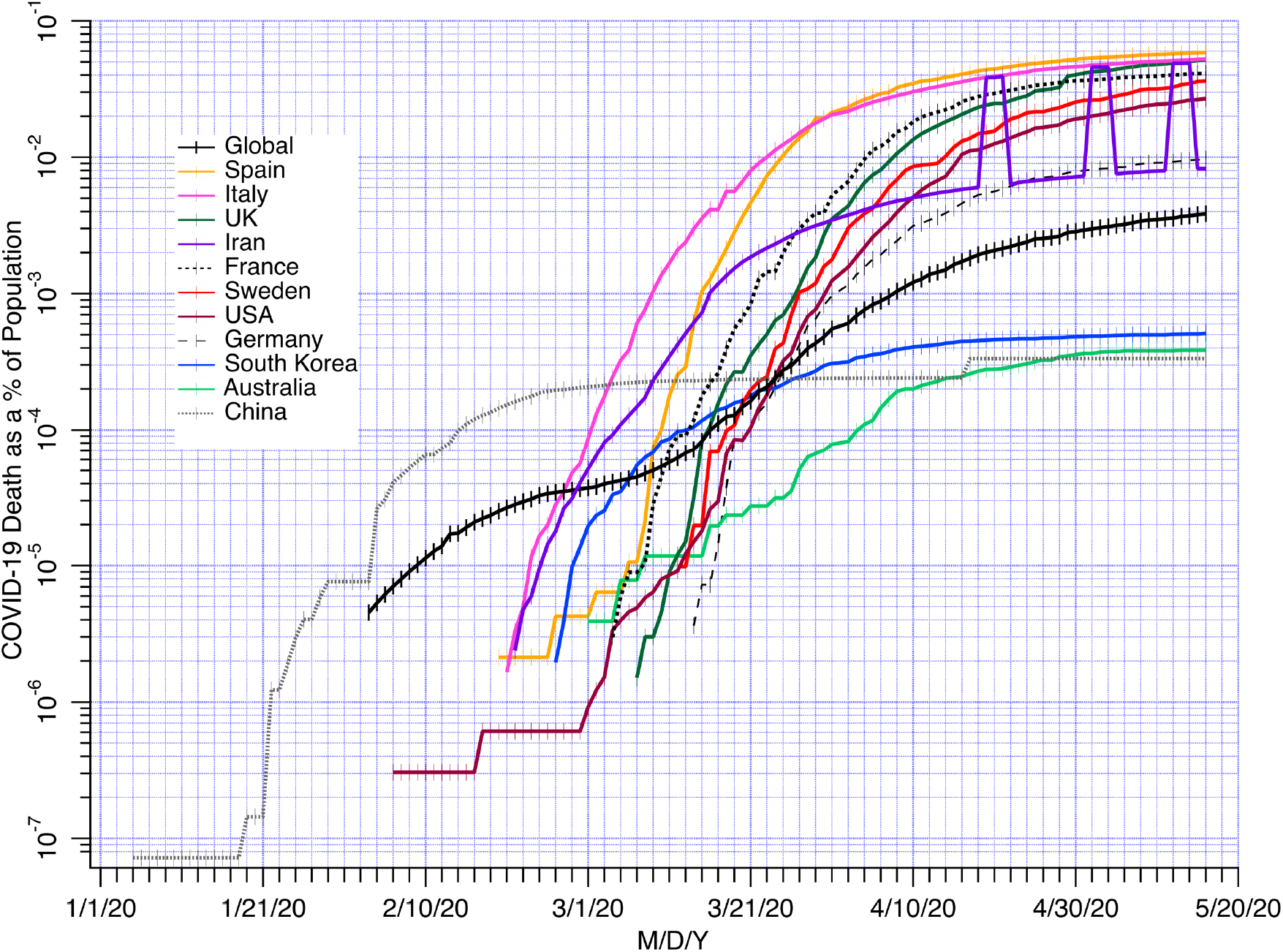
COVID-19 deaths as % of the population vs. time. Source: Center for Disease Control and Prevention CDC USA [4]. The minor ticks on the time axis each represent two days.

A secondary interest followed here is the statistics that could have led to the spacing by a mere 20 days of the mortality curves (measured at the same point) across many nations. To look at this properly we need to have either direct knowledge of the earliest cases outside of China (of which several are known) or as in the present approach a “Monte-Carlo” (dice-throwing) estimate of the size and frequency of gatherings combined with a known transmission probability for passing the infection from one who has a high titer of virus, to a non-infected contact at a gathering. In relation to gatherings, we have estimated that the number of days that one contacts a given number P_MAX_ of people is inversely proportional to P_MAX_. For example in author MWM’s 27,000 day life he once attended the Isle of Wight 1970 festival, with more than 100,000 people present, but daily he meets two or three people, as graphed in Figure 2. In between he has commuted daily on the London underground for three years, attended conferences, concerts, movies, parties of various scale, worked in small and large companies, and attended schools.

**Figure 2.**
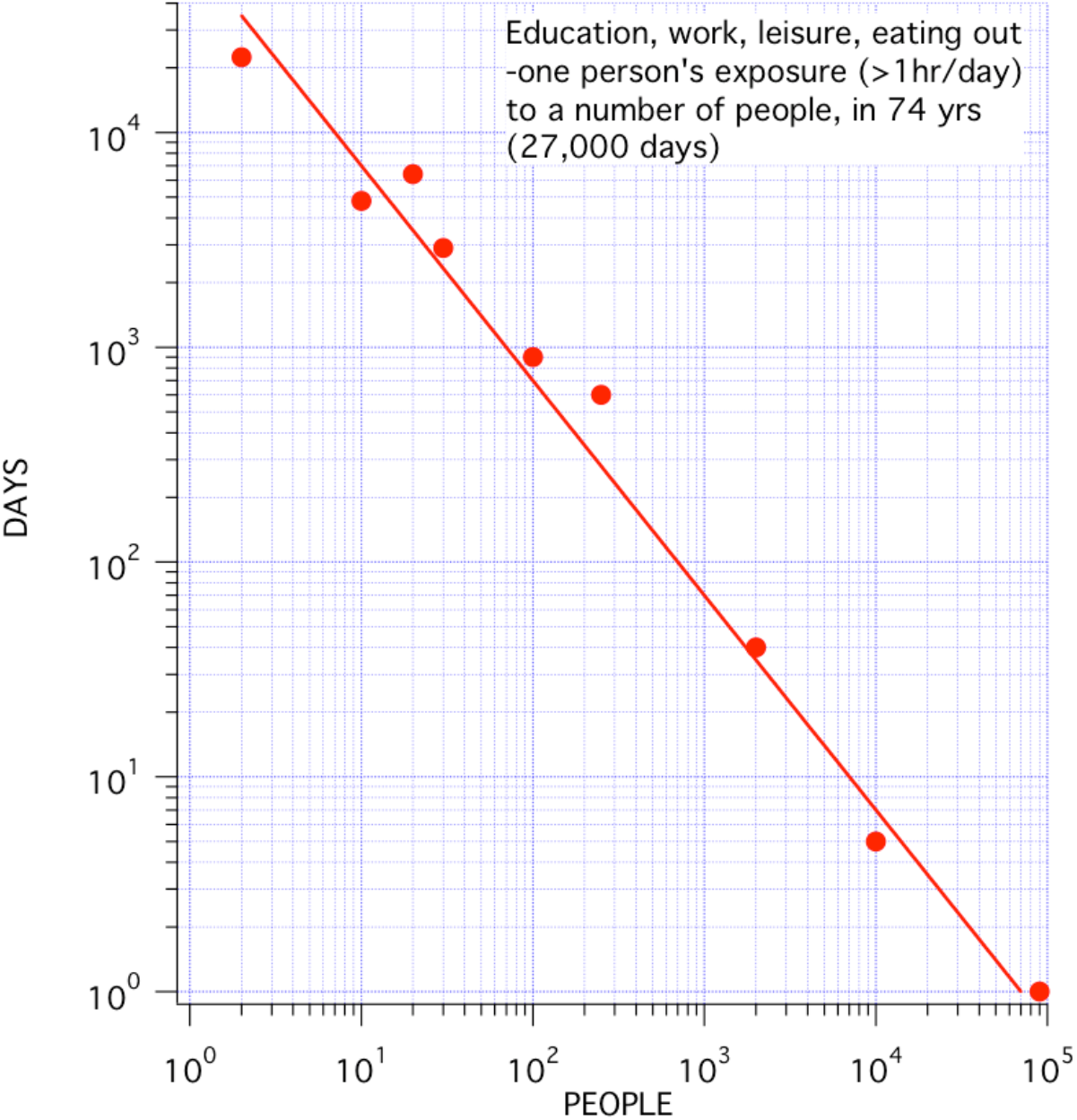
Illustrative example for one person of the (−1) power law relating number of days in contact with the specified number of people, summed through the day. The daily contact probabilities are found via division by 27,000.

## 2.1 Construction of the model: assumptions

The true situation is as complex as the number of the world’s population (excepting half of the identical twins) times the genomic range of the virus (there are currently about 200 mutations [5] but it is thought that to date none of these significantly affects its potency). It is standard practice in epidemiology to consider multiple levels of susceptibility to a pathogen. The simplest variation from a single level is two levels, the starting point of this analysis. Our two-state model makes the following assumptions:

1. There are at least two levels of susceptibiliity to attack by Covid-19. There will in reality be a full spectrum of response, but only two levels are needed to illustrate the effects of this internal variation. There may be many different reasons for this variation, further touched upon below.
2. The higher susceptibility is associated with a small fraction of the population.
3. The lower susceptibility is associated with the larger balance of the population.
4. The virus is constant in its genome.
5. The two levels of susceptibility are associated with effectively two probabilities (P_INF_) of transmission from an infected person to a person who has not previously been exposed. It is assumed that each of these levels is a constant per encounter, across all nations, at all times. (This constant, in a homogeneous population of <u>single</u> susceptibility, would be approximately 0.03, or 3%, to match the growth rate of the disease).
6. The course of the disease is assumed to be the same globally. This is taken here to be:
  a. 5 days incubation after the initial infection
  b. A 15 day course with symptoms, possibly slight, during which virus shedding reduces at the rate of 15% per day [6], a reduction factor of 0.85
  c. Either recovery, or death after 15 days with a fixed probability. The overall proportion of deaths in relation to cases is therefore taken as 0.85 to the power of 15 (= 0.087), which is the residual sick population after 15 days, times the probability of death (P_MD_) at day 15.
7. Once the patient has recovered, it is assumed that there will not be a recurrence, i.e. that person has entered what we denote in the program as IMPOP, the totally immune population.
8. Within a nation, in the low susceptibility subgroup there is an overall homogeneous level of contacts, ignoring the differences between city and countryside. This is probably the weakest assumption. It would be simple, if the gathering statistics were known for every locality, to make a larger model that adds all of them, and includes cross transfer via travel between them. Small sub-groups such as people on board ship, or essential transit system workers, have prolonged high contact rates that break the statistical trend for the overall population and put them in the higher susceptibility subgroup, even though their immune systems may be strong.
9. Within a nation there is a period of life as normal, in terms of contact rates and hygiene, during which the disease takes hold and enters exponential growth, followed by the realization that action has to be taken, and principally in that nation the size of gatherings is then dramatically curtailed (the process of “lockdown”).
10. In the sociology of nations, if one nation believes it is headed for the highest mortality it could be motivated to apply even more strict isolation so as not to be embarrassed by that honor.

## 2.2 The model: further details

The central entity in the model is an array of 20 numbers that represents the number of patients with a given viral load through the assumed 20-day progression of the disease. This array has placed in its first position the newly infected each day. Once per day (let’s say at midnight) the numbers are all passed up by one position, to leave the first position empty and ready to receive the next day’s new cases of infection. For the five days of incubation the number that is passed up does not change. At the beginning of the high viral titer on day 6, the number that is passed into the next array position 7 is reduced by 15% to reflect partial recoveries. This is viewed as a reduction to the number of patients, who otherwise, if still sick, continue to shed viruses at the same rate. As to the consequences leading to new infections, from day 6 onward in the progression array the infectivity stays at a constant level but since the number of sick is decreasing, the creation of new cases by these cohorts diminishes steadily.

This carries on for the next 14 days until the number entering array position 20 is reduced by 0.85 to the power of 15, to 0.087 times what it had been on initial entry to the array’s first position (day 1). At that point a mortality rate is applied with a value designed to align with experience.

In regard to the size of gatherings, each day every infectious case in that day’s cohort up to the number of 1,000 is allocated a “Monte Carlo” gathering size that is generated via inverse transform sampling [7], based upon an assumed (−1) power law for gathering size in normal circumstances. Above 1,000 an average contact number is assigned to the whole cohort to calculate the infections it creates. This process allows us to follow the start of an outbreak from single numbers, but reverts to higher computational efficiency once the number of cases rises to above one thousand in a cohort within the array. For convenient application of the results the standard population is taken as 10 million in all the data presented here. The mortality, or alternatively the infection numbers, can be converted to any one country’s population of N tens of millions by multiplying by N in direct proportion.

## 3. Model results for the “life as normal” case

### 3.1 Average contact numbers in normal circumstances

When the <u>maximum</u> number of daily contacts is limited to P_MAX_ the average number of contacts over a long period of days depends upon the distribution shown in Figure 2 and is obviously much less than P_MAX_. The sampling result for our assumed inverse first power law distribution (Figure 2) is shown in Figure 3.

**Figure 3.**
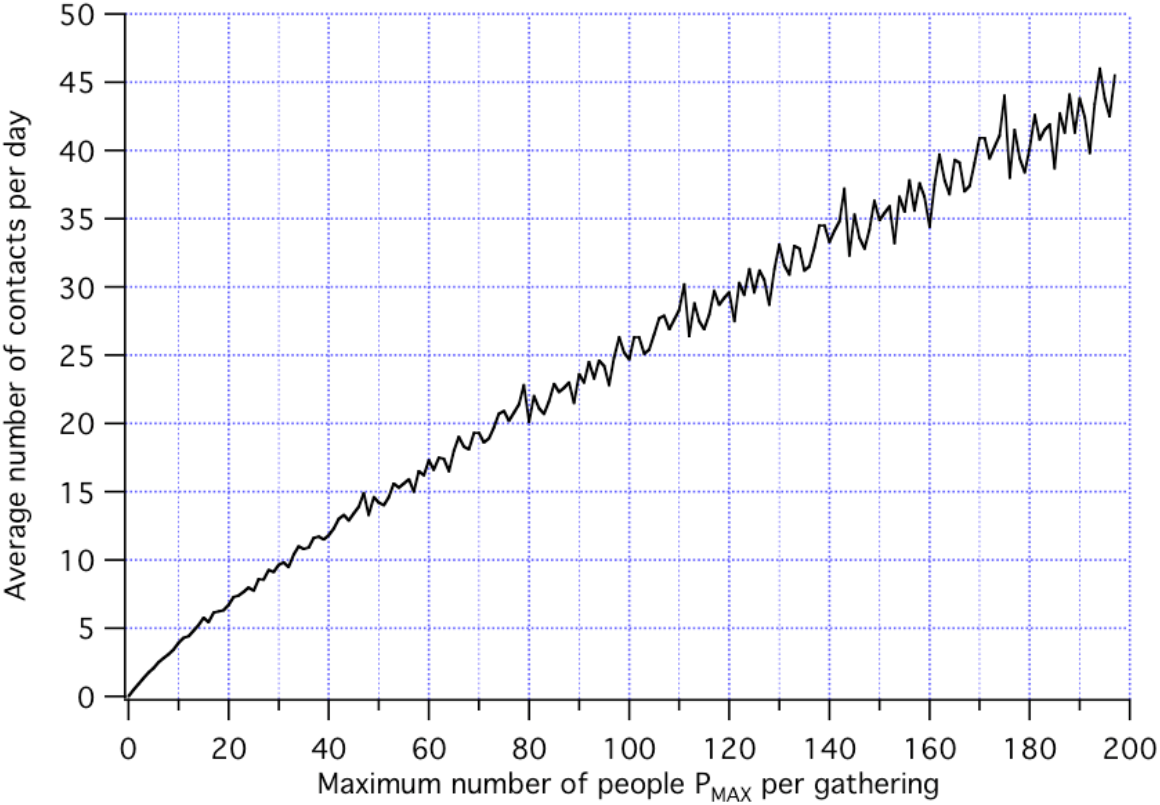
Average contacts per day versus a chosen cut-off maximum P_MAX_ of contacts, evaluated statistically from 1000 random samples of the inverse power distribution.

### 3.2 Results for a uniform population

To illustrate the difficulties with a uniform population and uniform susceptibility, a standard case was developed that matches the extremely rapid 3.1 day E-fold in the rising part of the curves in Figure 1. An E-fold is a 2.7x increase, which is the natural unit of exponential growth, representing an increase of unity in the exponent of E, the basis of natural logarithms. This E-fold number was derived from the slopes between 10^-5^ and 10^-3^ *%* mortality that were straight lines on the logarithmic plot of Figure 1. Countries such as China and S. Korea did not have a clear slope over two decades, due to rapid mitigation, hence their early growth rates, although comparably high, could not be well-established. According to the above assumption of a constant course for the disease once infected, the mortality growth rate in Table 1 applies also to the infection growth rate (in either a single or two-part population).

**Table 1.** Early growth rate of death due to Covid19 before introduction of mitigation, data in Figure 1.

| Nation | Days for 1 E-fold growth |
| --- | --- |
| Italy | 3.4 |
| Spain | 2.1 |
| UK | 2.7 |
| France | 3.1 |
| Sweden | 3.3 |
| USA | 3.7 |
| Germany | 3.6 |
| Average | $3.1 \pm 0.5$ (1 sigma) |

Figure 4 shows the modeled progression of infection and mortality with the growth rate of Table 1, in a uniform population. The disease rips through the whole population in 54 days, with 1.3 million new cases per day at its peak (in the standard 10 million population). A “life as normal” maximum daily contact rate of 200 was used, which gives an average daily contact rate of 45 (Figure 3). Clearly this picture does not remotely resemble the actual data. In particular:

1. Diagnosed active cases where the virus has been detected are up to 0.3% of the population, not up to more than 10% as this model would have it.
2. Infections have not dropped below 1 per day at day 54 in any nation.
3. Diagnosed cases have mortality between 5% and 15%, whereas the observed national mortality is less than 0.05%.

We also know that several different sectors of the population are disproportionately susceptible in most if not all nations:

1. The elderly, about 10% of a population, and especially those in care homes.
2. Poor people in crowded, stressful conditions, especially minorities within the USA.
3. Care givers, due to high exposure and inadequate protection.
4. (Still to be verified.) People taking medications tailored to the angiotensin system and its ACE2 enzyme that is the target of the SARS-CoV-2 spike protein.

We therefore need to depart from the uniform population model and move to the next simplest case: a two-part population.

**Figure 4.**
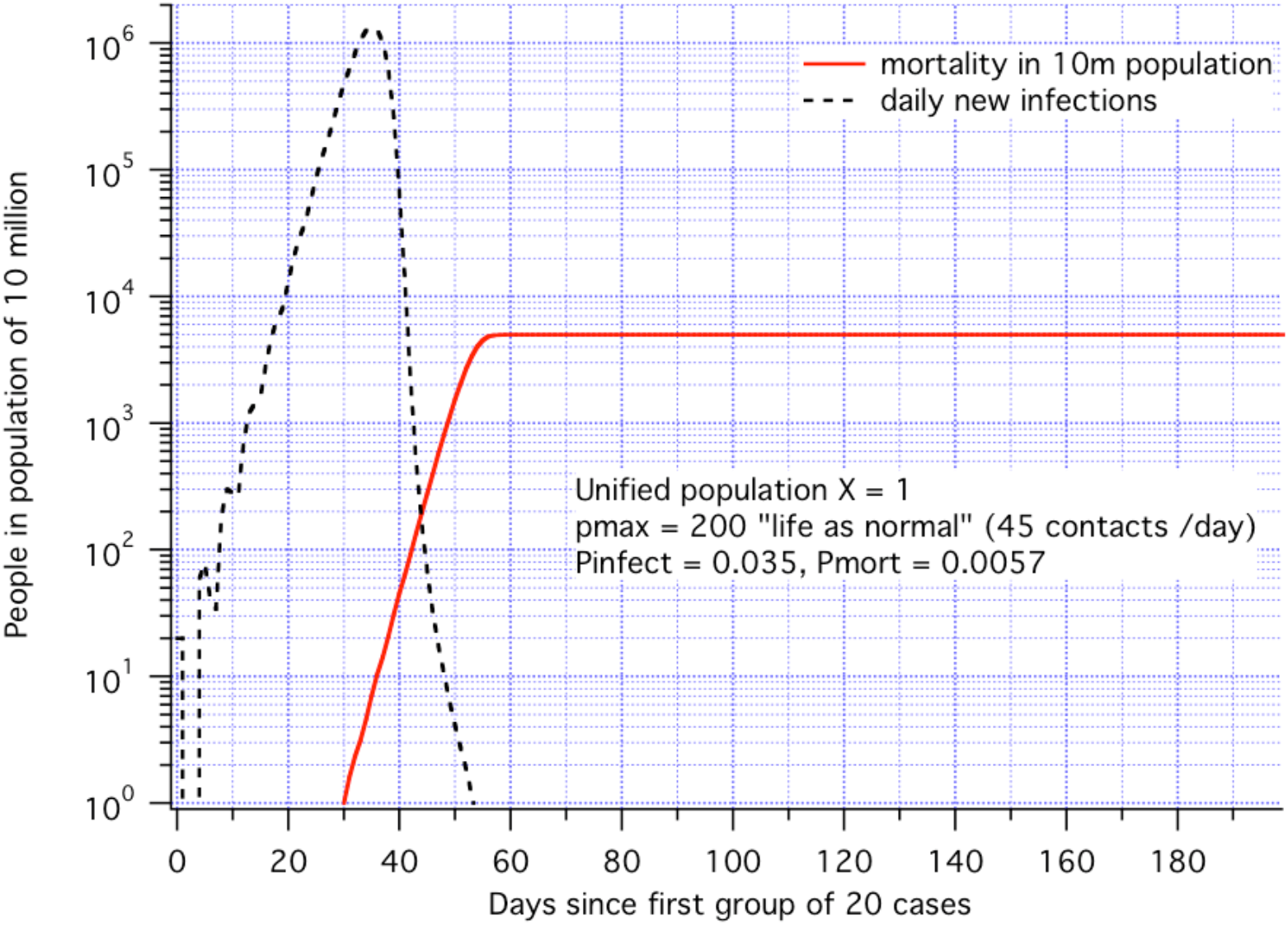
Propagation model for uniform population with E-fold = 3.3 days, matched to data. Mortality adjusted to give 0.05% plateau, population = 10^7^.

### 3.3 Results for a two-part population

We look at a population divided into two fractions with different susceptibility levels:

1. A higher susceptibility (vulnerable), fraction X of the population with high effective infection probability (P_INF_).
2. The balance (1-X) of the population having Y times less probability of infection (i.e. P_INF_/Y) whether due to youth, lack of confinement, or freedom from pre-existing medical conditions.

Clearly, many different sub-groups ought to be considered, and these will differ among nations, but here we only seek to understand the basic dynamics of the progression of Covid-19 within a two-sector population.

The results are very different from the single population case. For example, the disease will run through the vulnerable sub-population rapidly, tending to generate an initial plateau in deaths, but in the background it continues to spread more slowly through the larger, more resistant population.

Folded into this progression is the societal response, which varies greatly from nation to nation. One that takes aggressive measures to trace contacts and isolate cases reaches equilibrium at low mortality, but without most of the population having been exposed, therefore unable to benefit from “herd immunity”. This leaves sizeable residual un-infected populations in both the vulnerable and the high immunity groups, so that any new outbreak will have a short E-folding time and be more difficult to contain.

### 3.4 Two part population: the case of Sweden

Ideally we would compare the model to a nation that did not intervene at all, so the pure evolution of the disease would emerge. The next best resort is the country of Sweden, which did not impose a strict “lockdown”, however it did limit large gatherings. We jump right in with a two-step model for Sweden that we give the following trial parameters:

1. Initially “Life as normal” with maximum contacts P_MAX_ = 200, which gives an average daily contact number of 45 (Figure 3).
2. A 20% vulnerable fraction, i.e. X=0.2
3. A factor of ten less susceptibility in the (1-X) balance, i.e. Y=10
4. A probability of infection P_INF_ = 0.1 in the vulnerable population, i.e. P_INF_ = 0.01 in the low susceptibility population.
5. A transition on day 30 to partial social restrictions, i.e. to P_MAX_ = 50 with a daily average of 14 contacts (announced on 27^th^ March 2020).
6. A mortality of 0.01 x 0.087 to generate the 12^th^ May national statistic of 0.04%.
7. A one-day group of 20 cases on day 1, to pass through the random initial stage. These conditions gave the curves shown in Figure 5 for infections per day and fatalities.

**Figure 5.**
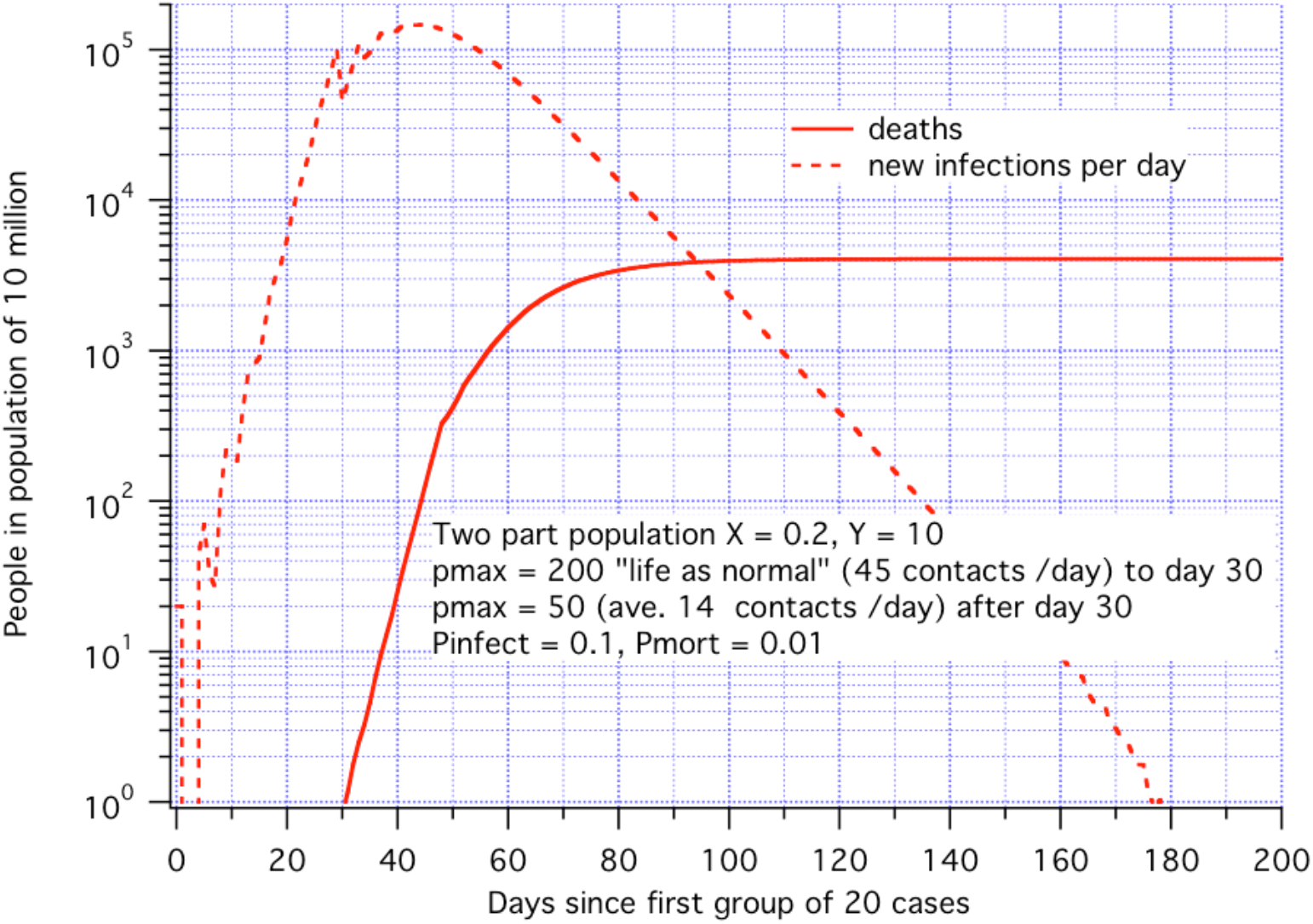
A model run of the two-part population relevant to Sweden. Partial social restrictions as defined were assumed to have begun on day 30.

The interesting finding is in Figure 6, which shows the fraction of each population that remains un-infected, vs. days into the Sweden model.

**Figure 6.**
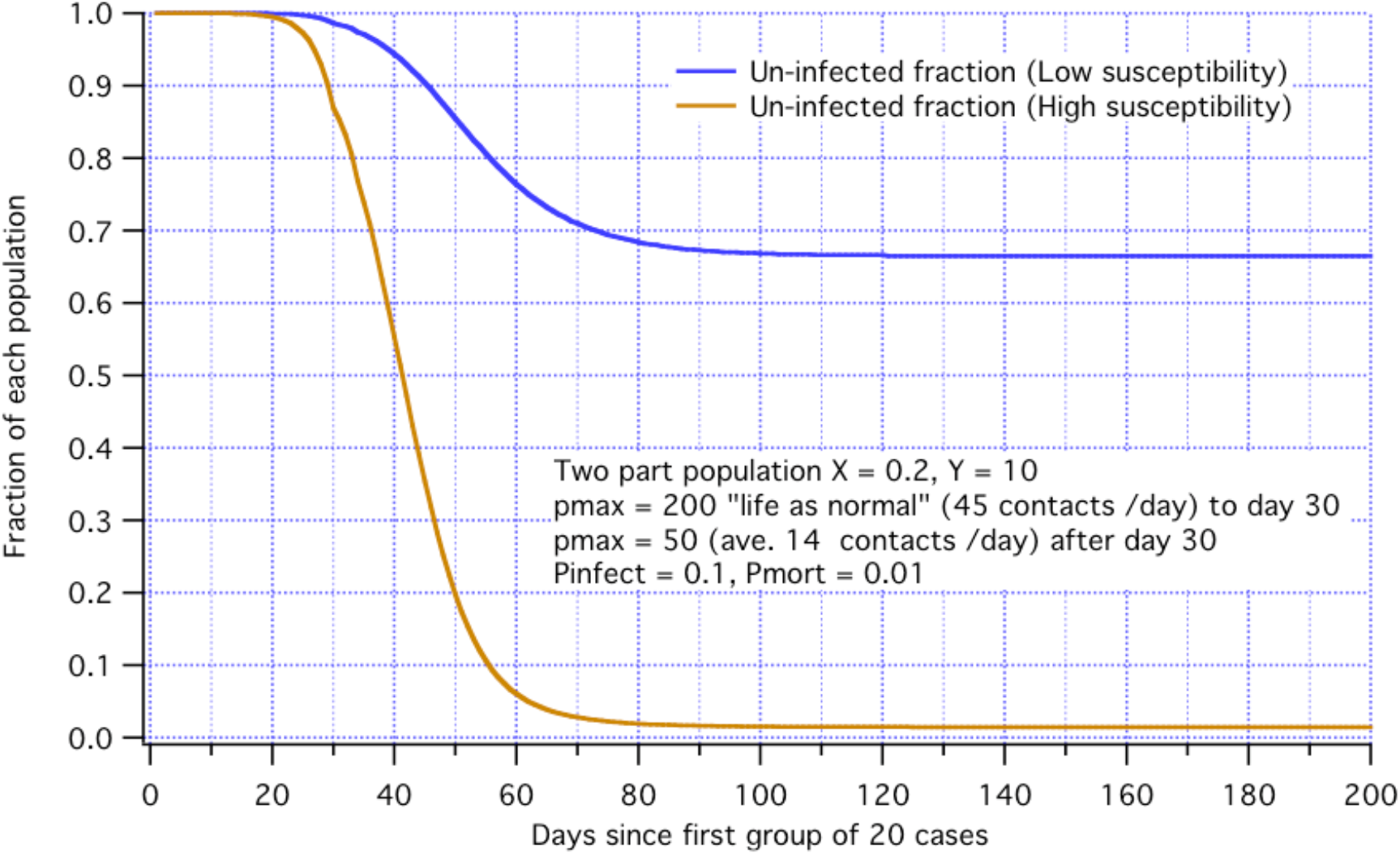
Fraction of each population that remains un-infected vs. days into the pandemic within a country such as Sweden with a relaxed social response on day 30.

Combining the infected fractions, about 50% of the whole population has been exposed by day 80, which is more than half the way toward “herd immunity”. The disease itself delivers new infections all the way out to day 177 (Figure 5), but the activity is insufficient to reach 66% of the low susceptibility population. For the parameters chosen, this therefore provides the de-facto definition of minimal herd immunity. This result is not to be taken too literally because it comes from an oversimplified picture. But it starts to show how the two-group model behaves. Ongoing data collection in Sweden [8] is beginning to suggest that many tens of percent of the whole population have antibodies to Covid-19 as of May12^th^.

### 3.5 Two part population: the case of China

China identified that Covid-19 had arisen in their population and was spreading rapidly [1,2]. What followed was a remarkable degree of decisiveness in which within days of the first 20 deaths there was imposed a rigorous “lockdown” in the city of Wuhan, and the Hubei province as a whole [9,10]. Starting with a baseline two-population model with “life as normal” P_MAX_ =200 we explore in Figure 7 the effect of the timing of hypothetical lockdowns. We start with timing because we find that the day the lockdown begins, relative to the onset of the disease, is much more important than the severity of the lockdown. In Figure 7 we consider a lockdown to P_MAX_ =10, corresponding to an average of 4 contacts per day (Figure 3) but we apply it at days 20, 25, 30 and 35 relative to the onset. The reported mortality in China is 0.0003% (Figure 1), or 30 in the 10 million population of our model, which places the China outcome between our day20 and day25 lockdown results. The Wuhan lockdown was applied on Jan 23^rd^ 2020 [9,10]. According to this model there would have been by that point approximately 3,000 infections per day in our model population of 10 million, many of them probably without symptoms.

With this early lockdown the estimated <u>un-infected</u> fraction of each of the two subpopulations remains close to 100% (Figure 8) in contrast to what we expect in Sweden. For the lockdown imposed on day 25 the fractions are 99% and 95% uninfected, in the two parts of the population. A later lockdown at day 35 yields 91% and 37% respectively. The early China lockdown would therefore not result in anything close to herd immunity.

As to the effect of lockdown severity, at a given day, for example day 30, reducing average contacts per day to 7, 4 and 2 would lead to deaths of 1300, 495 and 365 in the 10 million population modeled, indicating that little is gained via an extremely strict lockdown below P_MAX_ = 10, equivalent to 4 contacts per day.

**Figure 7.**
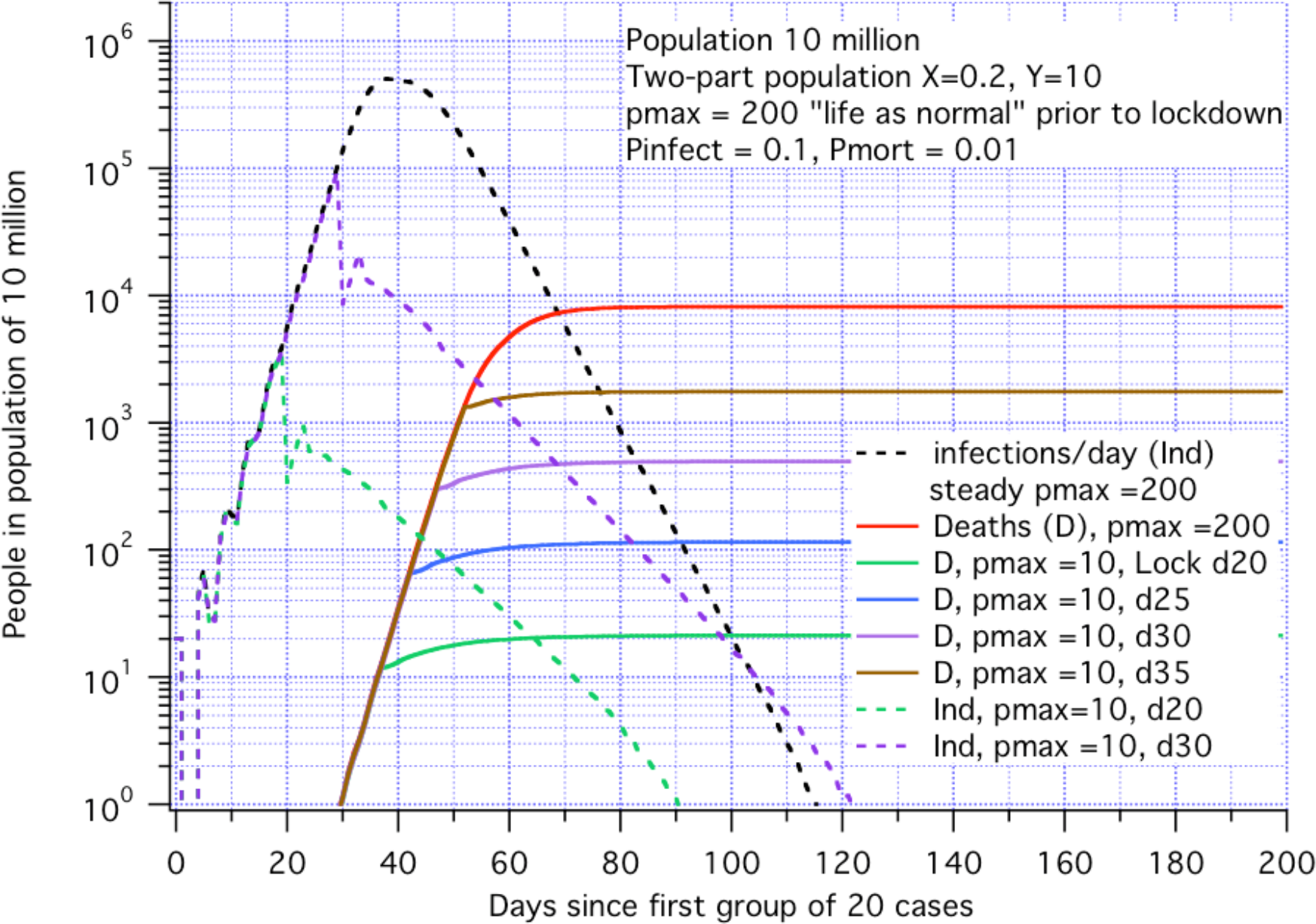
The baseline disease mortality profile (red line) with “life as normal”, and 4 lockdown cases, each to P_MAX_ = 10, but with different days (d) when applied.

**Figure 8.**
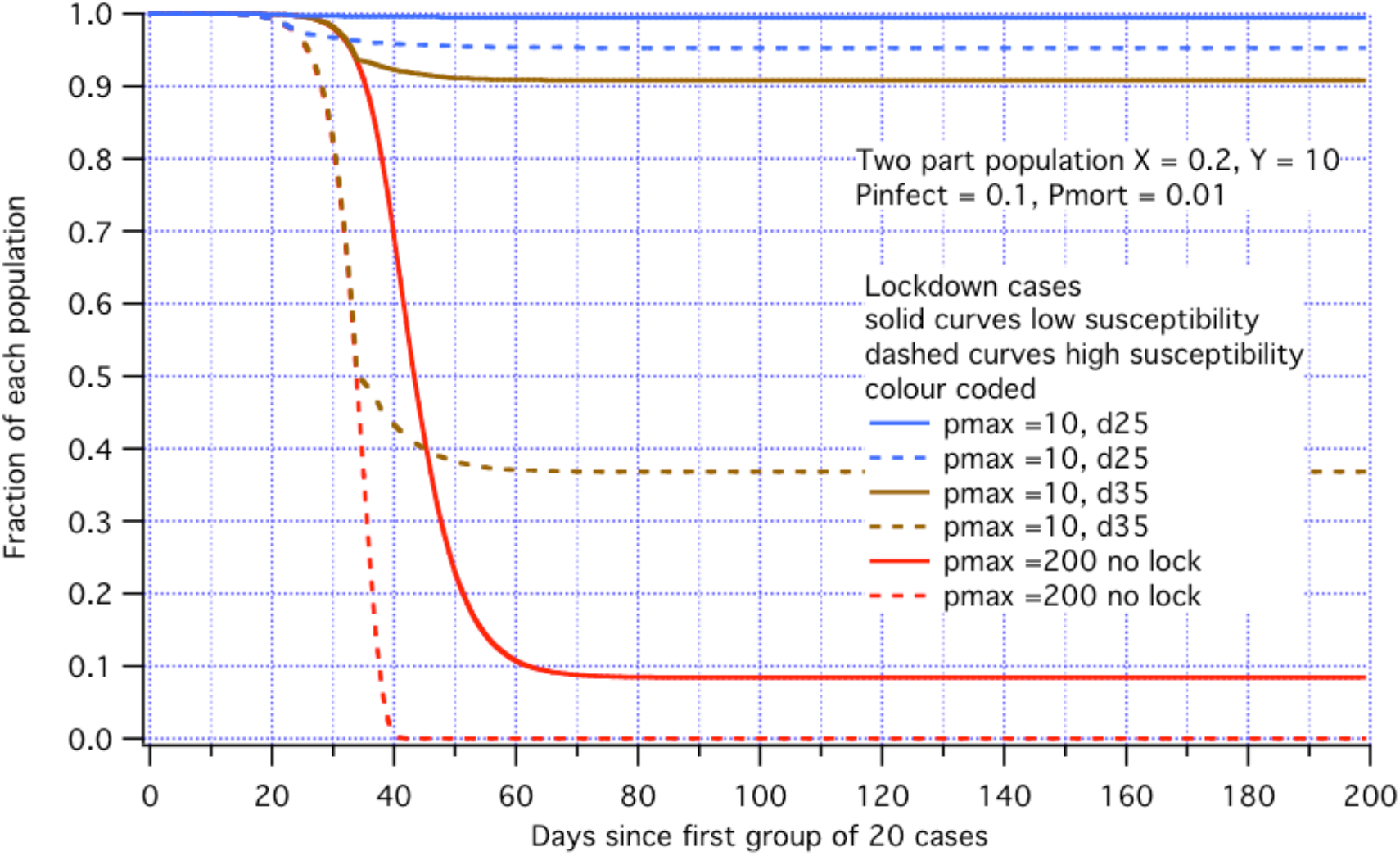
Fractions un-infected of low susceptibility (solid lines) and high susceptibility (dashed lines in two-sector population for: no lockdown; d25 and d35 lockdowns.

## 4. Discussion

The nature of Covid-19 is revealing itself as the pandemic unfolds. It does not cut through a population uniformly, but has greater effect on a relatively small fraction of the population. We identified mortality as possibly the most accurate measure of the progression of the disease in the absence of population-wide antibody data and have constructed a minimum model to fit the mortality data to date. As more information comes in on the fraction of people carrying antibodies to Covid-19, we hope to firm up the numbers relating to infection rates and immune fractions after infection, clarifying the approach to herd immunity (and what the criterion for that might be in this disease). It is possible that many countries are converging on a marginal level of herd immunity with a fractional death statistic of between 0.05% and 0.1% of the population.

It was assumed that the virus did not mutate significantly during the first 130 days of 2020 during which the mortality data of Figure 1 was collected that led to the model.

The world is becoming divided into one group of countries within reach of eradicating the disease, and another group of countries in which lax response (by design or otherwise) is leading inexorably toward universal presence of the disease and possibly herd immunity. This divide will inhibit travel and commerce between the groups.

Because the model is so “broad brush” and in an early stage of validation, it is not yet usable to predict the future in any one community or nation. Nevertheless, there is a striking consistency in mortality (as of 12^th^ May 2020) among the most affected group of nations, without any outlier on the high side, which begins to suggest that in this group an approach to herd immunity is well underway, and that it will take place without catastrophic additional mortality in the coming years.

## Data Availability

All data is available

